# A mixed methods exploratory study of family experiences of anticipatory injectable medicines at home: “It’s a really good idea but it’s just implementing it”

**DOI:** 10.1101/2022.06.29.22276695

**Authors:** C Faull, A Pilsworth, K McEvoy, H Hughes, L Bleazard, A Patterson

## Abstract

**Objectives:** To explore the experiences of bereaved family of anticipatory (JIC) injectable medications for symptom management and to identify ways to improve practice.

**Methods:** A single site, mixed methods study in two phases. A postal questionnaire sent to 100 family carers (FCs) achieved a 38% response rate of whom 79% volunteered for an interview and 14 were sampled to reflect use of medication, concerns and relationship with the deceased.

The descriptively analysed quantitative data and thematically analysed qualitative data were triangulated

**Results:** The vast majority (89%) perceived benefits to JIC medications but 18% of these described mixed feelings and issues, related to the context of dying and the responsibility for powerful medications. Communication was a key theme and some FCs described emotional impacts, misinterpretation of the intent of the medications and many desired more information to equip and empower them. There were diverse experiences of obtaining medications and guidance on storage and disposal resulting in distressing experiences.

Whilst most described good experiences of care and effectiveness of treatment 50% experienced delay in attendance of staff to administer medication. The majority of those interviewed would have been prepared to administer injections and the key driver for this was their experience of delays in resolving symptoms.

**Conclusions:** FCs were generally positive about JIC medicines and care received but they had also been a source of distress and anxiety and people wanted more information and empowerment. Professional support had often been difficult to access at the point at which it had been needed.

Key Messages

1. What was already known?
  - Anticipatory medications for symptom management in the last days of life are promoted as good practice by NICE
  - Health professionals support this but have identified complexities and concerns
  - There is no research exploring family or patient views and experiences
2. What are the new findings?
  - Family thought there were benefits but frequently described distressing experiences
  - Delay in support to administer medications was common
  - With appropriate supervision some family care givers would consider administration
3. What is their significance?
  a. Just-in-case medications are a trigger for thinking about dying. Practitioners need to anticipate that family may:
    - Realise for the first time that their loved one is going to die
    - Think that they are dying now, or more ill than previously suggested
    - Think these drugs actually end life.
  b. Family need more guidance about looking after a sick/dying person
  c. A PROM related to ‘time to attend’ should be considered.

## BACKGROUND

People that are dying will commonly experience one or more troublesome symptoms which may include pain, nausea and vomiting, delirium/agitation and respiratory secretions. If these symptoms are not managed effectively at home, patients and their families may suffer, may seek admission and may not fulfil their wish to die at home, [1-3]. Swallowing becomes problematic as people deteriorate and spend more time asleep or unconscious. Provision of injectable anticipatory or ‘just-in-case’ (JIC) medicines within patients’ homes, so that they are readily available to visiting health care professionals, is a key part of promptly managing symptoms and facilitating dying at home, [2-5]. Anticipatory prescribing is recommended in the National Institute of Health and Care excellence (NICE) guidelines on care of dying adults in the last days of life, [6] and the practice has become widespread [7,8]. There is published evidence detailing the experiences and the benefits and challenges identified by health professionals in utilising anticipatory medicines, [2,3,5,7-10].

The National End of Life Care Strategy (2008) encourages staff to engage with family and friends as co-workers in helping to manage a patient dying at home, [11] and whilst there is evidence surrounding the experience of these family/friend caregivers (FCs) managing the patient’s regular medicines and oral ‘as needed’ medication, there is little published evidence regarding the experience of FCs in relation to the JIC injectable medicines for use in the last days of life [7,12].

## AIMS AND OBJECTIVES

The study aim was to explore the experiences of bereaved FCs with regards to the provision and use of JIC medicines. The objectives were to:

- Understand the perceived benefits and concerns
- Identify good practice and barriers to good outcomes
- Make recommendations for improvements in care services and workforce training

## METHODS

### Design

A single site, mixed methods study in two phases designed to capture the broadest diversity of experiences. In phase 1 a postal questionnaire was sent to unselected FCs who met the inclusion criteria. In phase 2 a purposive sample of respondents to the questionnaire were interviewed to explore experiences in depth.

### Public and patient involvement

The focus of the research was suggested by FCs who had experienced challenges in the care of their loved one. The development of the study design, questionnaire and interview topic guide was supported through extensive consultation with bereaved FCs and the Hospice PPI consultee group.

### Participants

#### Phase 1

The hospice electronic patient record was used to identify 100 contactable FCs of consecutive deceased patients who had been prescribed JIC medicines on discharge from the inpatient unit or by the community specialist nurse. FCs, bereaved between 3 and 9 months previously, were sent a letter explaining the reason for contact together with a participant information sheet and questionnaire. One follow-up letter was sent.

#### Phase 2

Participants who completed the questionnaire were asked to indicate whether they would also be willing to take part in an approximately 1-hour, face-to-face interview with a researcher. Respondents were purposively sampled to include exploration of a diversity of experiences: those that had highlighted concerns and where practice had worked well. A diversity of relationships to the deceased was also sampled.

Both the questionnaire and interview sought information concerning FCs understanding and feelings about having the medicines at home; practical and process elements; experience of their use; and areas for improvement. Additionally, the appetite for administration of injectable medications by FCs themselves was explored in the interviews.

### Analysis

The categorical questionnaire data was analysed descriptively and free text responses analysed thematically. The interviews were audio recorded, transcribed verbatim and analysed thematically. Thematic analysis, used a constant comparison approach [13] and was developed through an iterative process of reflection, coding and research discussion. Analysis was undertaken alongside ongoing interviews allowing in-depth enquiry into emerging issues. Initial interviews were subject to open coding by two researchers (APi and APa). Emerging findings and development of the coding frame was discussed at project group meetings. Final coding of transcripts was undertaken by APi.

The sets were triangulated to build a composite picture of experiences and views. Quotes are presented in the findings by participant (P) number together with their provenance of questionnaire (Q) or interview (int).

## RESULTS

The response rate to the postal questionnaire was 38%. The characteristics of the sample are shown in Table 1. 64% of respondents were spouses, 63% of patients died at home and 82% had a cancer diagnosis. 97% (37/38) of respondents recalled having injectable medicines and analysis was performed on these 37 respondents. 65% (24/37) of people said that the JIC medications had been used, 27% said that they were not used and 8% were not sure.

**Table 1:** Characteristics of the participant sample and the relative that they cared for 76% of respondents indicated their agreement to be approached about an interview. We purposively sampled to include interviews of participants where they had indicated in the questionnaire that JIC medicines were not used (N=2), were used and there had been no problems (N=5), were used and problems had been reported (N=7). Our sample consisted of interviews with 17 participants related to 14 patients: 10 spouses (seven husbands, three wives), two daughters (two other daughters were interviewed with their parent), one sister and one cousin (one other cousin participant was interviewed with their relative). 9/14 primary interviewees lived in the same house as the patient, 50% were male and all were White British. Ten patients had died at home, three in a care home and one in LOROS Hospice. 71% had had cancer. The 14 patients ranged in age between 41 and 85 years.

|  |  | Number | % |
| --- | --- | --- | --- |
| Participant Gender | Male | 14 | 36.8 |
|  | Female | 24 | 63.2 |
| Relationship to deceased | Spouse | 25 | 65.8 |
|  | Daughter | 8 | 21 |
|  | Son | 1 | 2.6 |
|  | Sibling | 2 | 5.3 |
|  | Other relative or friend | 2 | 5.3 |
| Deceased Gender | Male | 21 | 55.3 |
|  | Female | 17 | 44.7 |
| Deceased Age | 40-50 | 2 | 5.3 |
|  | 51-60 | 7 | 18.4 |
|  | 61-70 | 9 | 23.7 |
|  | 71-80 | 10 | 26.3 |
|  | 81-90 | 10 | 26.3 |
| Deceased Ethnicity | White British | 34 | 89.5 |
|  | Indian | 2 | 5.3 |
|  | Other | 2 | 5.3 |
| Diagnosis of patient | Cancer | 31 | 81.6 |
|  | MND | 1 | 2.6 |
|  | Respiratory | 2 | 5.3 |
|  | Cardiac | 2 | 5.3 |
|  | Other | 2 | 5.3 |
| Place of Death | Home | 24 | 63.2 |
|  | Hospice | 9 | 23.7 |
|  | Care Home | 4 | 10.5 |
|  | Hospital | 1 | 2.6 |

### What did people feel about having JIC medication in the home?

89% (33/37) of respondents indicated that there were benefits to having anticipatory medications and 79% (26/33) felt entirely positive about having the medications in the house.

> *It is reassuring knowing that emergency medication for relief of symptoms is available without having the need for a doctor to visit to prescribe, then finding somewhere to get the drugs made up. All that’s required is getting the district nurse to administer*. P5Q

Perceived benefits orbited a number of themes

- Peace of mind.

> *It helped my mum’s anxiety knowing they were here if needed and also reassured me*. P37Q
- Quick and easy access to pain and symptom relief

> *They were first used in the middle of the night so were ready to use*. P24Q
- Reduced stress in obtaining drugs:

> *When I was originally given the prescription, the chemist didn’t have one of the items and couldn’t get it for a couple of days. It was reassuring to know it was available at my sister’s house when she needed it, instead of having to go around several chemists in an emergency*. P16Q
- Achieving patients’ wishes

> *It helped J stay at home longer and we were able to get married and then we went to LOROS for our ‘honeymoon’ for 14 days until his death*. P11Q

Despite perceiving benefits 18% (6/33)) described mixed feelings and issues related to having the JIC medications in the house. Thematically this mostly related to the context of death

> *Bit daunting as had to hide them away from my husband as he was in denial about his illness and upset me as I felt I wasn’t being honest with him and worried he might find them too. Brought home fact how bad things were and how close to death*. P30Q

but for some it was about responsibility for medications that were perceived, and sometimes communicated, as being powerful and potentially dangerous.

> *Having so many controlled drugs in the house is a huge responsibility for carers*. P19Q

Two respondents had only negative comments about JIC medications.

> *It was just horrible having the just in case meds. To top it all off once my husband passed away all the meds became waste*. P3Q

Another two respondents did not cite benefits and commented negatively about issues related to their use for symptom management.

### Information and communication about JIC medications

Most respondents recalled, but often vaguely, being given some information verbally but only one had had any written information. For some the medication had been ‘dropped off’ and explanation was given later when nurses came to attend the patient. A few said there had been no explanation.

Text comments summarised succinctly what was recalled by some and gives insight into communication between FCs and professionals and the impact of what is heard.

> *They were for emergency and in case the chemist was closed at the weekend and only to be used by the district nurse*. P15Q
>
> *The injections were always explained as “end of life” and were to help make the person comfortable in their final hours*. P31Q
>
> *We were told about each individual medicine and how/when they could/would be used. The reality of this was a little daunting as it confirmed advanced fears of the future but also helped us to understand what could be used when the time came*. P23Q

The interviews give more insight in to what lies behind the recalled communication and contextualises such dialogue. Several spoke of how discussing the time when the JIC medications might be needed really brought home the reality that their loved one was dying.

> *I was sort of staring in the abyss and I thought, you know, what is going to happen, how am I going to be able to manage this. I had not realised the full implications of the medications I had not been told (that she was dying) at that time you see*. P1int

The terminology used was somewhat problematic. The term ‘end-of-life’ medication was frequently recalled. For some this was seen in retrospect as a euphemism and didn’t convey clearly enough that the patient was close to dying, whilst for others it conveyed that the patient was close to death which was at odds with what they had heard from clinicians. For some the term ‘end-of-life’ conveyed that the medications were to end life or hasten dying

> *I know there was more than one drug there and but there must have been*.
>
> *I am assuming there is an end-of-life injection*. P18Int

Another theme in communication was the context of risk and safety seen in both in the text responses to the questionnaire and in the interviews.

> *She said “don’t worry about them we will sort them out when the time comes”. You don’t touch these because they are like they are …*.. *they are a deadly cocktail aren’t they*. P21int
>
> *The need to keep them under lock and key was explained but this was impracticable. With my children being older, I was not concerned*. P34int

About 50% of respondents to the questionnaire would have welcomed written information although there were some hesitancies voiced.

> *It would have been helpful to see written information on each medicine for the family, but for the person dying I’m not so sure. It would depend on how they felt about dying*. P23Q

Emphasis was clear that verbal information was paramount, together with the opportunity to ask questions and that written information was an additional source of reference, something to refer back to and share with others in the family. The interviews identified that such information sharing was broader than just detail of the medication, involving contextualising of such discussions of when they might be needed, what problems the patient might be experiencing and equipping the FC to assess and act effectively.

However, 38% said that they would not want written information and 19% were unsure. Two key reasons for this emerged: that they had sufficient explanation verbally and that it wasn’t something of relevance to their role. Some comments were especially thought-provoking concerning the burden on FCs that written information and the expectations that this can convey.

> *I had enough to deal with other tablets and the circumstances I was in. I didn’t need any more information on something I had no control over*. P4Q

### Obtaining and storing JIC medication

Almost 10% of people reported significant issues (days and longer) in obtaining the medications from a pharmacist. For some this increased their appreciation of the anticipatory stocking-up, for others it was an additional stress.

> *So I had to go to three different ones and he was at home all the time really poorly. …….And he was almost frightened of being on his own because he said I don’t want to die when you are not here* P11int

In one case, the need for restocking of the injectable medications, led to the FC not being with their wife when she died.

There was little consistency in how risk, responsibility and requirements for safe storage was approached by professionals. There was an apparent fine balance to be struck between sufficient and insufficient emphasis on this issue. Some FCs recalled this as quite frightening and/or that their integrity seemed questioned.

> *Well if you find something missing the police will be involved* P13int

Others were given prescriptive information on storage and were told that they had to be in a locked box. Some FCs reported that they would like secure arrangements for the medications for fears of them going missing and then consequently being blamed.

Some FCs were given no information and stored them with other oral medications and some expressed concerns.

> *They literally sat in the lounge with him. I think that’s quite loose, shall we say, for controlled drugs to just be sitting around in somebody’s home. There was quite a variety of scenarios that could happen with them*. P25int
>
> *Because it wasn’t in a locked container, I didn’t know how trustworthy they [paid carers] were or whether it was open for abuse*. P5int

For some FCs the information given by professionals that the drugs were controlled, added to their burden

> *As for me and the wife two elderly people only in the house then I didn’t feel too bad about it. Obviously kept my fingers crossed that none went missing*. P18int

Some FCs worried that their home was a potential target to be broken into or that paid carers might take medications that they were responsible for.

> “*Because you know people around know what is going on don’t they and they know that somebody is in that state, they could quite easily break in to your property and get the drugs. Because people do nowadays they are desperate aren’t they. So, it is a huge responsibility I think having those tablets and injections and things in your house*. P19int

The risk, responsibility and requirements for safe storage was encountered as a problem in pharmacies by two FC interviewees.

> *And one wouldn’t let me have the medication because I didn’t have proof of a locked cabinet*. P11int

### What happened when JIC medications were needed and used?

Medications were reported as used mostly for pain and distress or agitation. Text comments in the questionnaire mostly described good experiences of care and effectiveness of treatment.

> *My partner was in tremendous pain in the early hours, so being on my own I dialled 111 and a doctor came out and gave him an injection, which helped*. P17Q
>
> *As his pain increased the nurse gave the injection first then as the pain and distress worsened the syringe driver was fitted. I believe this was all given at the correct time, as my father deteriorated, he was never left in pain or distress for long. I just rang and a nurse came out within 20/30 minutes*. P35Q

Interviewees also identified excellent care with well trained staff whose timely visits made them feel supported and safe.

> *Nurse said I think you ought to ring and he came (Doctor) within quarter of an hour*. P1int

Others recalled that whilst staff provided excellent care, the patients and staff were let down by the system

> *I think I had to ring twice and when A (Nurse) got to me she said that she apologised for the delay but the call had been put through to a different team. And she just happened to spot S’s (Patient) name in the call queue*. P25int
>
> A*nd I rang again and the nurse said I will be there probably about an hour but she had got another call locally to us to a nursing home. And she didn’t actually get to us until about half past two in the morning during which all of that time (4 hours) D had been shouting*. P9int

Nearly half of FCs answering the questionnaire identified that deciding when to call for help was a key concern (Table 2) and although the majority knew who to call for help around a third of people didn’t. These did not emerge as distinct themes in the interviews, rather that they were aspects of the bigger picture of how challenging it can be to care for a dying loved one and the many responsibilities that FCs feel.

**Table 2.**
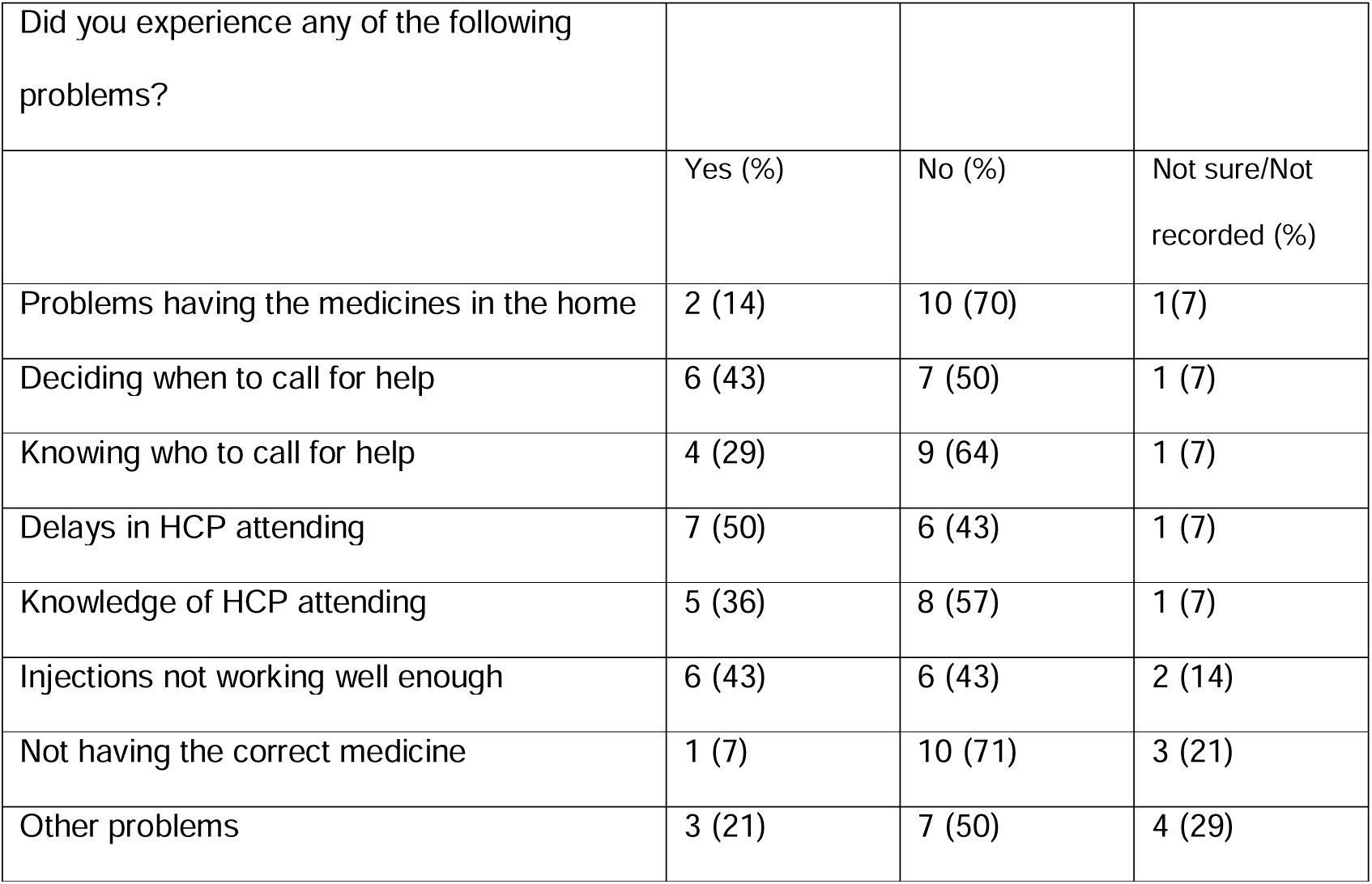
The problems that FCs reported where patients had used JIC medications

### Problematic care

Over half of FCs however encountered at least one problem (table 2). 50% of FCs responding to the questionnaire experienced delay in attendance of staff to administer medication. The in-depth interviews identified that this delay was sometimes a considerable number of hours. It appeared to be most noted when outside of ‘normal’ working hours of services and was very problematic when patients were moved between places of care.

> *Sometimes it would take 3 hours to get a nurse, especially in the middle of the night*. P1int
>
> *Having extreme pain and having to wait an unknown time and it can be for hours and it was say two hours probably when they got to us on one occasion. But never straight away and of course 15 minutes is 15 minutes too long*. P26int

FCs called for help when a loved one was in distress and the emotional impact of this waiting was substantial.

> *We were led to believe that that wouldn’t happen. It’s going to be horrible, it’s not going to be nice; but you will look after her with drugs to make it as comfortable as possible. We are just watching out mum in agony and desperate and actually asking us to help her*. P8int
>
> *That was the worst bit [waiting for symptom relief], through everything*.
>
> P9int

In some instances, staff who attended were not able to give medication. This was sometimes because they did not have the knowledge or skills and commonly it was ‘*that bit of paper’*, authorisations to administer was not in place.

> . *I called the emergency people out. I had got the paperwork and she said “it’s not been signed off” …… So I said “does that mean you can’t give it?” and she said “no I can’t”*. P18int

Some HCP’s lacked confidence in decision-making and administering JIC medications and end-of-life care. Some interviewees recounted instances of crying by professionals faced with difficult decisions or patients dying.

> *The [care home] matron on the phone was crying almost because they were telling her at the other end of the line to get a grip of herself as good as, that was the gist of it. It was terrible*. P1int

This added to the relative’s burden at a difficult time

> *And he (The Carer) actually stood in the bedroom one day crying at the state my dad was in. I didn’t quite know how to deal with him (the carer) really because you are trying your best not to be a mess yourself ……it was quite hard for me* P19int

### How the experience might have been improved

There were many comments expressing gratitude and satisfaction.

> *I don’t know how it could have been improved. If the medicines were not here, I don’t think I could have looked after him at home. For that I am extremely grateful. That is what J wanted. To stay at home*. P18Q

The comments of some respondents were very emotive and provide significant comment of quality of services and the burden and impact of FCs.

> *Nightmare of watching my wife in extreme agony. Everyone concerned did their best, but the system is overloaded*. P1Q
>
> *On the whole care professionals gave an excellent service, however the time spent alone with the patient was lengthy and this was very distressing at times when the patient was in pain and suffered a lot of agitation*. P19Q

People asked for improvements both directly and indirectly by telling of their regrets and distress. A few were specific to JIC medication but most were related to end-of-life experience more broadly [Box 1 below].

The absence of instructions about what to do with medications after the person had died and the challenges they experienced in this was frequently mentioned and many were concerned about the waste. For some this issue was of significant importance relating to the experience of grief and loss.

> *And I did think of topping myself with it. I took it to our local pharmacy and they wouldn’t take it. That was when I thought I’ve had enough. I’m going to take it all. I had to get it out of the house*. P11int

#### Box 1

**FC reflections on what needs to be improved**

##### Specific to JIC medications

- *Clarity and provision in arrangements for return/disposal of drugs*
- *Just to ensure people attending are aware of what, when and how much to give and if not sure to seek assistance*.
- *Quicker response by on call nurses*
- *Medicines in stock!*

##### End-of-life care more broadly

- *Obtaining and collection of equipment*
- *My biggest regret is that she was unable to spend her end-of-life with her wonderful friends and staff at the hospice*.
- *Waiting hours for a private ambulance was difficult. He was being taken back to the hospice and we felt helpless*.
- *The only problem at a time of great stress having to fight for assistance from care services and I struggled for a while*
- *I didn’t know what to expect and I think more info on what action to take for certain symptoms and when to call for help versus severity of things. I just got on with it felt a bit alone in it and hard to ask for help*.
- *More information would have helped. I had no experience of looking after a sick person. The nursing help, doctor and carers were there but I never knew how ill my husband was. I’m still not sure I did everything right and because of the medication on his last day I never got to say goodbye*.

### Appetite for administering themselves

When asked in the interview (n=13) 10 of the FCs suggested that they would have administered injectable medication themselves, albeit somewhat reluctantly, if they had been taught the skills and given guidance and support. Two others said that someone else in their family would have been prepared to do this. A key driver for this was the pain or distress of a loved one.

> *Probably, if it had come to it, I would probably have done it. Yes, there is no two ways, I would have had to just suck it up and do it*. P6int

Delays in getting trained staff appeared to increase their motivation to do this and those with prior experience of giving injections for other reasons or those of clinical backgrounds felt confident and perhaps disempowered not to be supported in this.

> *I have no problem injecting insulin and organising tablets and oramorph and all that. So looking back you see I was told it has to be a nurse and I was never given the option of I would you be able to do it. Considering the number of things they have trained me up to do I was, that was something I could have done without a doubt*. P26int

The need for legal clarity and safeguards was identified by participants but not everyone agreed with this involvement for themselves or others.

> *One, I would go against training somebody to do that at home and two, I would be against it…*.. *putting a deadly substance …*..*totally against it*. P13int

Some felt it was not within their capability and the potential emotional impact featured highly in whether they would be prepared to do this.

> *It would be quite difficult to know when to give the medication. If she dies tomorrow – Christ, I would never forgive myself*. P8int

## DISCUSSION

This is the first study that specifically focusses on FCs experiences of JIC anticipatory injectable medications. Our findings indicate that the vast majority were reassured by their presence. However, despite seeing the overall benefits overall, almost one in five FCs described mixed feelings and for a few they caused considerable distress. The down-side of JIC medications was sometimes related to feelings of responsibility for medications that were perceived, and sometimes communicated, as powerful and potentially dangerous. But more often these feelings of distress related to the JIC being a harbinger of dying. For a few the distress this caused was very great indeed.

The considerable effort and burden of responsibility for medications, especially controlled drugs, has been described by Pollock *et al* in the recent in-depth study of FCs managing medicines at home [12]. Our work expands on this in exploring how injectable medications and patients’ needs in the last days of life can increase this feeling of responsibility. This burden and the related anxieties seem to be heightened further by the attitudes of health professionals to the risks and the lack of clarity in how such medications need to be stored and monitored in the home and later accounted for and disposed of. Our findings align with others that indicate this is an area that needs improvement to minimise the burden for FCs and provide proportionate guidance for the safety of all parties [12].

Experiences of the use of JIC medications were, in the most part, of good care with effective and responsive symptom management but some participants described poor experiences where HCP knowledge seemed to be inadequate. Others have identified that community nurses can find decision-making about ‘as required’ injectable medications challenging and tend to err on the side of caution [3,9,14]. Most often however experiences of poor care in this study were related to system failure. Several respondents described having to witness the distress of their loved-one when the systems/services failed to provide timely support to manage symptoms. Unlike many emergencies in health care there is no service standard that is defined for speed of response; in this case attendance and administration of medication that has been provided and authorised. Our findings indicate that the time to respond to symptom distress is pivotal, suggesting that patients/FC are failed when attendance takes longer than an hour and very satisfied when this is within 30mins. These findings indicate the need to operationalise the 4^th^ Ambition for end-of-life care (Care is coordinated: *I get the right help at the right time from the right people*),[15]. In an area of care that has few patient-related outcome measures, a clear standard for this would be valuable.

A few accounts described a transition of care to hospice, hospital or care home, which was usually precipitated by the difficulty of achieving symptom management at home. These were times when care might deteriorate further and FCs recalled that their knowledge and experience was ignored in the context of hospital and care home, but not in hospice; this is far removed from the family as co-worker aspiration of the end-of-life care strategy [11] and echoes the findings of others [12,16].

Communication about the rationale for JIC medications was pivotal to FCs although their needs varied greatly as to the detail they desired. The implication for practice is that information needs to be tailored, purposeful, timely and mindful that this may be the first time someone realises their loved one is dying. It seems important to ask about what these medications convey to them and correct any misperceptions that they may have. Our findings also suggest that it is important to actively clarify that the purpose of the medications is not to hasten death or end life (a perception that patients and FCs may not share with professionals) but that they are used to maintain comfort in the same way as tablets medications. Written information was considered useful but not a replacement for verbal discussion which offered opportunity for questions as well as education/empowerment that some people wanted about deterioration and dying concerning what would happen, what to watch out for and what to do. The prescription and dispensing of JIC could provide an ideal opportunity for this work and could be another standard for end-of-life care. However, it has been observed that prescribing JIC ‘early’, well before deterioration is occurring, may mean perhaps that professionals don’t undertake this level of communication, which doesn’t seem pertinent to either party at that time [5,12]. The pharmacists role in enhancing explanations and empowerment could potentially be explored.[16-18]

A high proportion of our interviewees would consider extending their role as co-workers to undertaking administration of injectable medications to manage symptoms. The greatest drive to this was waiting for professional attendance and witnessing “*agony”* and *“distress”* of loved ones. A number of the interviewees had experience of injecting medications for other health needs, commonly insulin and heparin, which gave them technical confidence and a number were of health or care backgrounds affording them situational judgement and decision-making confidence. However, FCs articulated the emotional burden this may have for them and some felt overwhelmed by the prospect of taking on greater responsibility for care, echoing others’ findings [12,19]. This role is established in other countries and has been cautiously advocated in the UK during the COVID pandemic but the evidence base remains poor to ensure safety of all parties [20, 21]

The mixed methods approach has promoted breadth in recruitment however participants were required to be able to read the English questionnaire. The response rate was 38% which is considered high for a post-bereavement questionnaire on a sensitive topic. Although perhaps those with strong views about their experiences (both positive and negative) may have been more likely to respond than others, the diversity of experiences recalled in the interviews would seem to argue against that. Participants all had some contact with specialist palliative care services so their experiences might differ from those that do not. The single research site may identify findings unique to this context including the significance of the system issues which may be less or more impactful elsewhere.

## CONCLUSION

Most FCGs found having anticipatory medicines generally beneficial Carers describe distressing experiences especially where there has been poor communication and timely professional support has been difficult to access. A good experience relies heavily on the timeliness, confidence and competence of staff and accessibility of services. Empowerment of FCs and more clarity on storage, monitoring and disposal of JIC medications is called for. Seeing a loved-one suffering with symptoms was the key driver for consideration by some family care givers of administration of injectable medications under advice/supervision but they identified potential emotional consequences to this.

### Implications for practice

1. Mindful, purposive, timely and tailored communication:
  - This may be the first time someone realises their loved one is dying
  - They may perceive that they are dying now/more ill than people have said
  - They may think these are drugs to end life
2. Use the opportunity of JIC medication planning to educate and empower people about what to expect and when to call for help
3. Be clear about storage and what to do with unused drugs after death

### Implications for services

- A ‘within 1 hour’ standard for attendance time would provide a good outcome
- Develop a carer administration protocol/SOP for the few people that would feel they would like to do this.

### Implications more broadly

- Minimum standards and best practice guidance for storage, monitoring and accounting for medications needs to be developed.

## Data Availability

All data produced in the present study are available upon reasonable request to the authors

## ACKNOWLEDGMENTS

Design of the study and protocol and document development Claire Wilkins and Sam Krauze. PPI input to shaping the research question, design of the questionnaire and topic guide for interviews Monica Glover and Elaine Dolman. LOROS research support Emma Bowler, Natalie Ayton and Wendy Gamble. Participants who so generously gave of their time and emotional energy

## ETHICAL APPROVAL

favourable ethical opinion was given by the Leicester South Research Ethics committee in May 2019 ref 19/EM/0049.

